# Survival Outcomes With LUCAS-Assisted vs Manual CPR in In-Hospital Cardiac Arrest Obese Patients

**DOI:** 10.1101/2025.10.19.25338320

**Authors:** Aayushi Pareek, Ivan Huespe, Sinner Jorge, Aryan Pathak, Saurabh Sujanyal, Said Bateh, Jose Delgado, Anek Jena, Raksha Venkatesan, Vrutti Patel, Deeksha Thatigolla, Ananya Biswas, Faiz Saleem, Titilope Olanipekun, Abby Hanson, Jeff Smith, Karen Stoner, Jose Soto Soto, Anna Shapiro, Parag Patel, Devang Sanghavi, Pablo Moreno Franco, Nick Kelly

## Abstract

**Background:** To compare 24-hour and 60-day survival after in-hospital cardiac arrest in overweight (BMI ≥ 25 kg/m^2^) and obese patients (BMI ≥ 30 kg/m^2^) resuscitated with LUCAS® assisted versus manual CPR, and to determine whether BMI modifies this association.

**Methods:** Multicenter retrospective cohort of adult IHCA events at seven Mayo Clinic hospitals from January 2019 to January 2025. Cardiac arrests <4 minutes or without a BMI record were excluded. IPTW adjusted for demographics, arrest location, and comorbidities. We first compared 24-hour and 60-day mortality between groups using IPTW-weighted logistic regression. To assess effect modification by BMI, we fitted analogous models with restricted cubic splines (knots at 25, 28, 37 kg/m^2^) and LUCAS® × BMI interaction terms. The model choice was based on Akaike Information Criterion.

**Results:** A total of 679 patients were included (manual CPR=595 and LUCAS® assisted=84). In weighted analyses, no statistically significant difference was observed in 24-hour mortality (OR 1.54; 95% CI: 0.94–2.53; p = 0.09) or 60-day mortality (OR 1.83; 95% CI: 0.92–3.64; p = 0.09). Spline models showed that 24-hour and 60 days mortality increased modestly between BMI 25-28 kg/m^2^ and then plateaued above 28 kg/m^2^ in both groups. No LUCAS® × BMI interaction was significant (all p > 0.14), indicating consistent effects across the BMI spectrum.

**Conclusion:** In overweight and obese IHCA patients, LUCAS®-assisted mechanical CPR did not improve 24-hour or 60-day mortality versus high quality manual compressions, and effectiveness was independent of BMI.

## Introduction

Obesity is a growing global health concern that not only increases the risk of mortality but also complicates the clinical management of critically ill patients, particularly during cardiopulmonary resuscitation (CPR). In-hospital cardiac arrest (IHCA) is a major contributor to inpatient mortality, and obesity has been associated with worse outcomes in critical care settings due to impaired chest wall compliance, difficulty in achieving effective manual compressions [1].

The use of mechanical CPR devices, such as the Lund University Cardiopulmonary Assist System (LUCAS®) has offered a promising alternative to traditional manual chest compressions. These devices aim to deliver automated compressions at a consistent depth and rate, with active decompression, potentially overcoming limitations of manual CPR in patients with challenging body habitus. [2,3]. The American Heart Association (AHA) guidelines have acknowledged the utility of mechanical devices like LUCAS®, particularly in environments where manual CPR may be compromised, such as during transport [4].

Despite the growing use of mechanical CPR systems, comparative survival data for obese patients with inhospital cardiac arrest remain scarce. This gap is clinically important because excess body mass and altered chest-wall mechanics can impair delivery of highquality manual compressions [5]; mechanical devices could therefore plausibly improve outcomes in this population. Previous trials have shown feasibility and improved hemodynamic profiles associated with mechanical compressions, yet these findings are not specific to obese populations and are often limited to out-of-hospital settings [2,6]. The impact of mechanical CPR on post-resuscitation outcomes, particularly 60-day mortality, remains underexplored in obese patients with IHCA.

This study aims to fill a critical evidence gap by evaluating whether mechanical CPR improves survival outcomes in overweight and obese IHCA patients, a population increasingly encountered in clinical practice.

## Material and methods

### Study Design and Setting

We conducted a multicenter, retrospective cohort study across the seven Mayo Clinic hospitals; these hospitals had a combined bed capacity exceeding 4,000. All consecutive adult cardiac-arrest events that occurred between 1 January 2019 and 30 January 2025 were screened using electronic health record review. The data was accessed on March 15, 2025, and the authors had access to information that could identify individual participants during or after data collection. Patients were followed from the moment of cardiac arrest until death, hospital discharge, or 60 days post-arrest, whichever came first. This study has Mayo Clinic Institutional Review Board approval (ID#:24-012976) and was reported in accordance with the Strengthening the Reporting of Observational Studies in Epidemiology (STROBE) guidelines. As this study was retrospective in nature, the requirement for informed consent was waived by the institutional ethics committee.

### Participants

Eligible individuals were patients aged ≥ 18 years, who experienced an in-hospital cardiac arrest managed at any Mayo Clinic facility, and had a documented body mass index (BMI) recorded within the preceding 365 days. For the primary analysis, we restricted the cohort to patients with BMI > 25 kg/m^2^, because our objective was to evaluate the effectiveness of the LUCAS® device in overweight and obese patients. We excluded cases lacking a valid BMI entry, cardiac arrest date, or arrests that ended within four minutes, as these patients didn’t have sufficient time for LUCAS® placement.

### Variables and data sources

We extracted patient demographics (age, sex, selfreported race/ethnicity) and anthropometrics (height, weight, and BMI) from the electronic health record. Preexisting comorbidities, including hypertension, diabetes mellitus, chronic kidney disease, chronic obstructive pulmonary disease, heart failure, coronary artery disease, and prior stroke, were identified via ICD10–CM codes recorded before arrest. Cardiac arrest characteristics (arrest location within the hospital - intensive care unit vs. general ward—and total duration of chest compressions) were abstracted from standardized resuscitation flow sheets in the electronic health record.

The exposure of interest was the mode of chest compression used during the index resuscitation. “LUCAS® CPR” was defined as chest compressions that could start manually but switched to uninterrupted compressions delivered exclusively with a LUCAS®™ 2 or LUCAS®™ 3 device from its application until either return of spontaneous circulation (ROSC) or termination of efforts. “Manual CPR” was defined as compressions provided entirely by human rescuers without the aid of any mechanical device. Data was extracted from the resuscitation flow sheets from the electronic health record.

Two pre-specified outcomes were evaluated, including: 1) Early mortality was defined as death within the first 24 hours after arrest, and 2) In-hospital mortality encompassing all-cause death occurring during the same hospitalization, up to a maximum of 60 days after the arrest. Vital status and time of death were obtained from discharge summaries and institutional death registries.

### Manual CPR

Upon recognition of inhospital cardiac arrest, manual chest compressions were initiated immediately by Advanced Cardiac Life Support certified providers. These clinicians undergo quarterly highfidelity simulation training that incorporates realtime audiovisual feedback to monitor and optimize compression quality ensuring a rate of 100–120 compressions per minute, a depth of 5–6 cm, and full chest recoil in accordance with American Heart Association guidelines.

### LUCAS®-Assisted CPR

Patients who received mechanical compressions began with manual compressions as above, but the LUCAS®™ device was applied as soon as practicable typically within the first two to three minutes of arrest recognition. After device placement, compressions were delivered at a fixed rate of 102 ± 2 per minute and a depth of 5.3 cm with active decompression on recoil. Rhythm checks were performed every two minutes during brief device pauses, following standard Advanced Cardiac Life Support procedures.

### Statistical analysis

Baseline characteristics were summarized as mean ± standard deviation or median (interquartile range), depending on normality assessed by the Shapiro-Wilk test. Categorical variables were reported as counts and percentages. Between group comparisons (LUCAS® vs. manual CPR) employed two-sample ttests or Mann-Whitney U tests for continuous variables and χ^2^ or Fisher’s exact tests for categorical variables, as appropriate.

To adjust for measured confounding, we applied inverseprobabilityoftreatment weighting (IPTW) using stabilized weights from a logistic regression propensityscore model. Covariates included patient race, sex, arrest location (ICU vs. general ward), and the following comorbidities: diabetes mellitus, chronic kidney disease, chronic obstructive pulmonary disease, congestive heart failure, peripheral vascular disease, cerebrovascular disease, pepticulcer disease, and mild or moderatetosevere liver disease. We assessed covariate balance by comparing standardized mean differences (SMDs) before and after weighting, defining adequate balance as all postweighting SMDs < 0.10. To minimize the influence of extreme weights, we truncated any weight exceeding the 99th percentile at the 99thpercentile value [7].

In the IPTWweighted cohort of patients with BMI ≥ 25 kg/m^2^, we fitted logistic regression models for two endpoints (24hour and 60day mortality. First, we estimated the overall association between LUCAS® use and each mortality outcome. Second, to assess whether this association varied across the excessweight spectrum, we modeled BMI in two ways: (1) as a linear continuous term and (2) with restricted cubic splines (knots at the 10th, 50th, and 90th percentiles: 25, 28, and 37 kg/m^2^) to capture potential nonlinearity. In both specifications, we included the main effect of LUCAS® and an interaction between LUCAS® and the BMI terms, either the linear term or the spline basis functions to test for effect modification. We compared model fit using the Akaike Information Criterion (AIC) and selected the specification with the lowest AIC. From the chosen model, we report odds ratios and 95 % confidence intervals for LUCAS® use and for BMIspecific effects. All statistical analyses were performed with STATA v.16.

## Results

Of 883 inhospital cardiac arrests among patients with BMI ≥ 25 kg/m^2^, we excluded 55 events (6.2 %) lasting < 4 minutes and 149 events (16.9 %) lacking a BMI measurement within the 365 days before arrest, yielding 679 eligible cases: 595 (87.6 %) received manual CPR and 84 (12.4 %) received LUCAS® assisted CPR (Fig 1). Baseline characteristics are presented in Table 1. The cohort’s mean age was 66.3 ± 13.6 years, mean BMI 29.9 ± 4.9 kg/m^2^, and 32.4 % were female. The most common comorbidities were heart failure (25.0 %), chronic kidney disease (17.4 %), diabetes without complications (13.1 %), and prior myocardial infarction (11.5 %).

**Table 1.** Demographic data.

|  | All (N=679) | Manual CPR<br>(N=595) | LUCAS® CPR<br>(N=84) | p.overall |
| --- | --- | --- | --- | --- |
| Female, n (%) | 220 (32.4%) | 189 (31.8%) | 31 (36.9%) | 0.413 |
| **Race, n (%):** |  |  |  | 0.077 |
| White | 565 (83.2%) | 490 (82.4%) | 75 (89.3%) |  |
| Black/African American | 61 (8.98%) | 59 (9.92%) | 2 (2.38%) |  |
| Asian | 23 (3.39%) | 21 (3.53%) | 2 (2.38%) |  |
| Other / Unknown | 30 (4.42%) | 25 (4.20%) | 5 (5.95%) |  |
| Age, Mean (SD) | 66.3 (13.6) | 66.4 (13.7) | 65.5 (13.3) | 0.586 |
| BMI, Mean (SD) | 29.9 (4.90) | 30.0 (4.97) | 29.4 (4.32) | 0.29 |
| SOFA day before CPR, mean [IQR] | 3.00 [1.00;5.00] | 3.00 [1.00;5.00] | 2.00 [1.00;4.00] | 0.018 |
| Minutes CPR, mean [IQR] | 23.0 [10.0;41.0] | 22.0 [10.0;41.0] | 25.0 [11.5;41.2] | 0.584 |
| ICU, n (%) | 176 (25.9%) | 161 (27.1%) | 15 (17.9%) | 0.095 |
| **Comorbidities, n (%):** |  |  |  |  |
| MI | 78 (11.5%) | 64 (10.8%) | 14 (16.7%) | 0.159 |
| CHF | 170 (25.0%) | 150 (25.2%) | 20 (23.8%) | 0.886 |
| PVD | 68 (10.0%) | 63 (10.6%) | 5 (5.95%) | 0.258 |
| Cerebrovascular disease | 24 (3.53%) | 23 (3.87%) | 1 (1.19%) | 0.343 |
| COPD | 72 (10.6%) | 62 (10.4%) | 10 (11.9%) | 0.822 |
| Rheumatic disease | 19 (2.80%) | 15 (2.52%) | 4 (4.76%) | 0.278 |
| Diabetes (no complications) | 89 (13.1%) | 80 (13.4%) | 9 (10.7%) | 0.602 |
| Diabetes (with complications) | 35 (5.15%) | 28 (4.71%) | 7 (8.33%) | 0.182 |
| Renal disease | 118 (17.4%) | 105 (17.6%) | 13 (15.5%) | 0.736 |
| Cancer | 65 (9.57%) | 61 (10.3%) | 4 (4.76%) | 0.161 |
| Severe liver disease | 24 (3.53%) | 21 (3.53%) | 3 (3.57%) | 1 |

**Fig 1.**
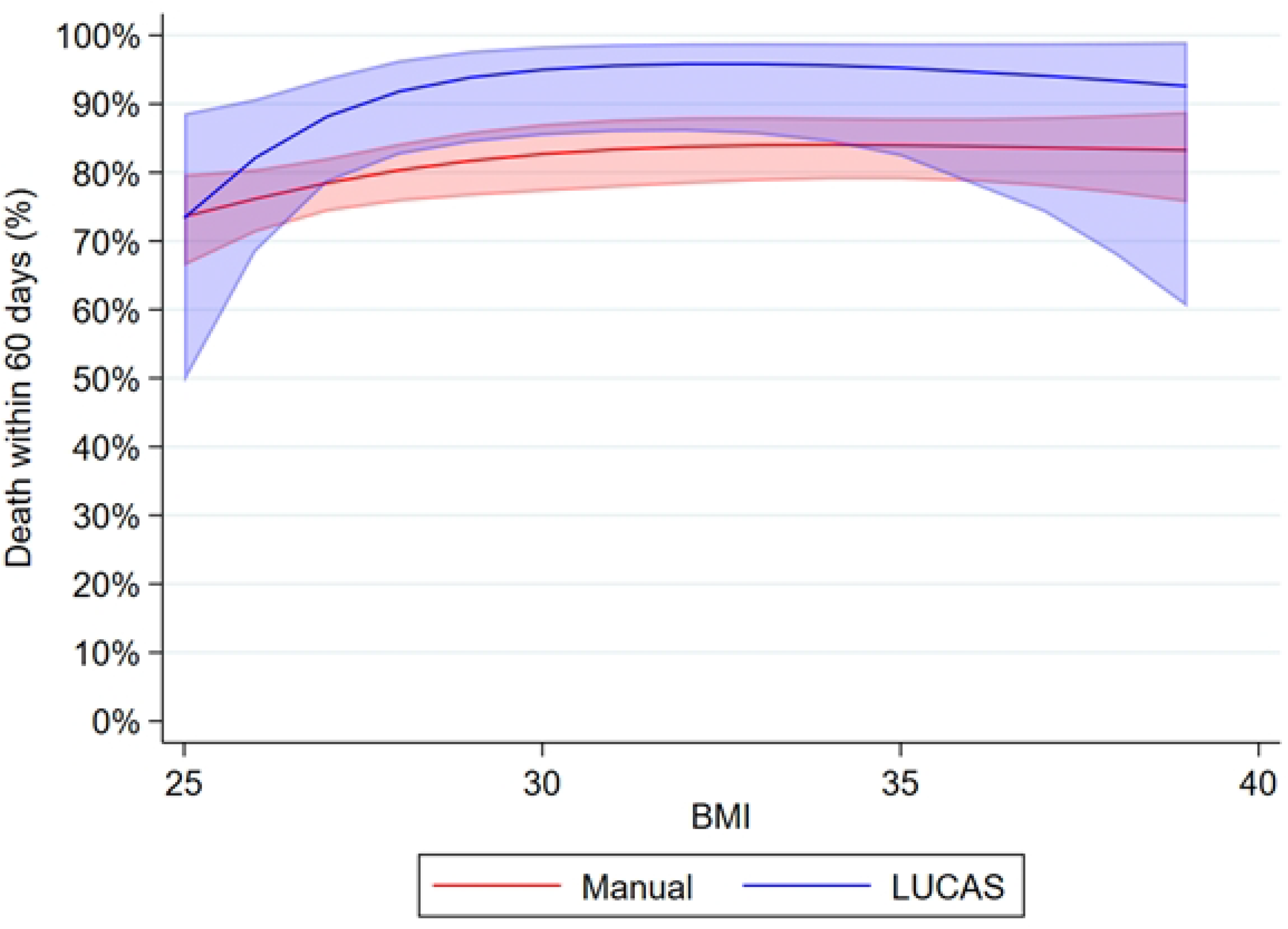
Flow diagram illustrating exclusions and final cohort allocation for patients with BMI ≥ 25 kg/m^2^ experiencing in-hospital cardiac arrest.

### Mortality within 24 hours

IPTW reduced every standardized mean difference to < 0.10, indicating good balance between patients who received manual CPR and LUCAS® CPR (S1 Fig). Among patients with BMI > 25 kg/m^2^, crude analysis showed no significant association between LUCAS® use and 24-hour mortality (OR 1.47, 95% CI: 0.92-2.33; p = 0.11), and the analysis in the IPTW-weighted subpopulation showed that LUCAS use was associated with a non-significant trend toward increased mortality with an OR 1.54 (95% CI: 0.94-2.53; p = 0.09).

Across the BMI spectrum, the restricted cubic-spline model fit the data better than the linear specification (AIC 1919 vs. 1920) and was therefore retained. In this model, the LUCAS® main effect (log-odds −1.70; 95% CI −11.37 to 7.97; p=0.73) did not differ from the intercept, indicating no baseline difference at BMI 25 kg/m^2^. The first spline segment (BMI 25–28 kg/m^2^) had a modest but statistically significant positive coefficient (0.12; 95% CI 0.01 to 0.24; p=0.04), consistent with a small rise in 24-hour mortality as BMI increases within this range (Table 2). The second segment (BMI 28–37 kg/m^2^) was negative but non-significant (−0.18; 95% CI −0.41 to 0.05; p=0.12), suggesting a risk plateau beyond BMI 28 kg/m^2^. In practical terms: the modest increase in mortality between BMI 25–28 kg/m^2^ suggests a transitional risk zone, while the plateau beyond BMI 28 kg/m^2^ indicates stabilization of risk irrespective of CPR method. Importantly, neither spline×LUCAS® interaction was significant, meaning the BMI–mortality curve had a similar shape for both manual and LUCAS®-assisted compressions. Predicted 24-hour mortality risk curves with 95% CIs (Fig 2).

**Table 2.** Coefficients derived from the restricted cubic spline models for both outcomes (mortality within 24 hours and 60 days mortality)

| **Death within 24 hours** | **Coef. (95 % CI)** |
| --- | --- |
| LUCAS® CPR | −1.70 (−11.37 to 7.97) |
| Spline 1 (BMI 25–28 kg/m<sup>2</sup>) | 0.12 (0.01 to 0.24) |
| Spline 2 (BMI 28–37 kg/m<sup>2</sup>) | −0.18 (−0.41 to 0.05) |
| Spline 1 × LUCAS® | 0.08 (−0.28 to 0.44) |
| Spline 2 × LUCAS® | −0.17 (−0.87 to 0.53) |
| Intercept | −3.35 (−6.48 to −0.22) |
| **Death within 60 days** | **Coef. (95 % CI)** |
| LUCAS® CPR | −9.53 (−23.09 to 4.02) |
| Spline 1 (BMI 25–28 kg m<sup>−2</sup>) | 0.14 (−0.01 to 0.29) |
| Spline 2 (BMI 28–37 kg m<sup>−2</sup>) | −0.21 (−0.50 to 0.09) |
| Spline 1 × LUCAS® | 0.38 (−0.13 to 0.89) |
| Spline 2 × LUCAS® | −0.65 (−1.62 to 0.33) |
| Intercept | −2.50 (−6.52 to 1.53) |

**Fig 2.**
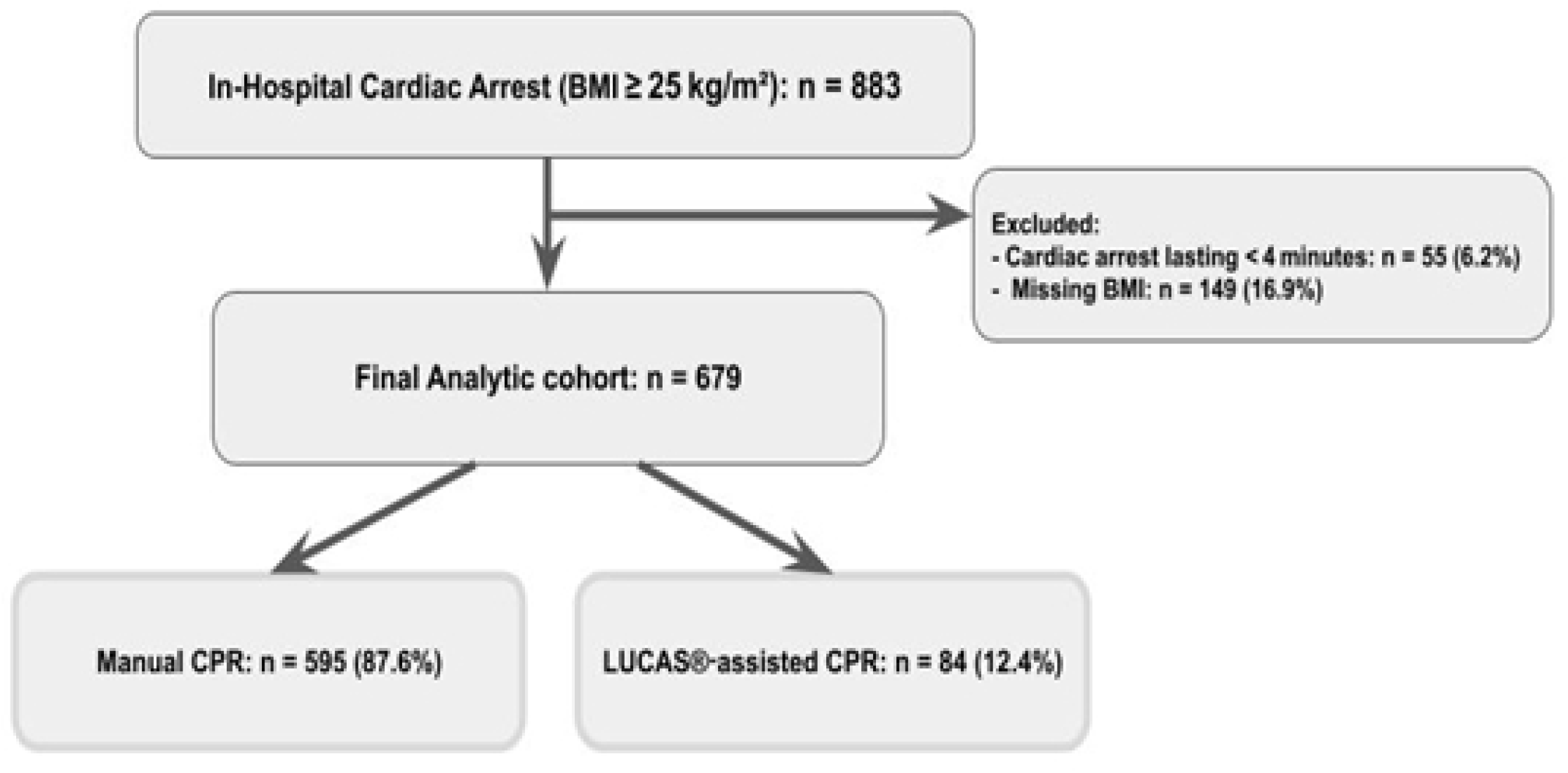
Predicted probability of death within 24 hours after in-hospital cardiac arrest across the BMI spectrum (≥ 25 kg m^−2^) Curves derive from the IPTW-weighted cubic-spline logistic model. The red line depicts manual CPR and the blue line LUCAS® CPR; shaded bands indicate 95% confidence intervals.

### Mortality within 60 days

In the unadjusted cohort of patients with BMI ≥ 25 kg/m^2^, LUCAS® use was associated with a nonsignificant increase in 60-day mortality (OR 1.69; 95 % CI 0.87–3.29; p = 0.12). The IPTW-weighted analysis yielded a similar estimate (OR 1.83; 95 % CI: 0.92 to 3.64; p = 0.09), indicating that adjustment for measured confounders did not materially alter the result.

To assess whether the association varied across BMI, we compared a linear model with a restricted cubic-spline specification; the spline fit better (AIC 1245 vs 1249) and was retained (Table 2). In this model, the LUCAS® main effect (log-odds −9.53; 95% CI −23.09 to 4.03; p=0.17) was not significant at the reference BMI of 25 kg/m^2^, indicating no baseline benefit or harm. The first spline segment (BMI 25–28 kg/m^2^) showed a small positive coefficient (0.14; 95% CI −0.01 to 0.29; p=0.07), whereas the second segment (BMI 28–37 kg/m^2^) was negative but non-significant (−0.21; 95% CI −0.50 to 0.09; p=0.17). Taken together, the modest rise in risk between BMI 25– 28 kg/m^2^ suggests a transitional risk zone, while the leveling above 28 kg/m^2^ indicates a plateau irrespective of CPR method. Neither spline×LUCAS® interaction was significant (p=0.14 and 0.19), showing that the BMI–mortality curve was essentially the same for manual and LUCAS®-assisted CPR. Predicted 60-day risk curves with 95% CIs (Fig 3).

**Fig 3.**
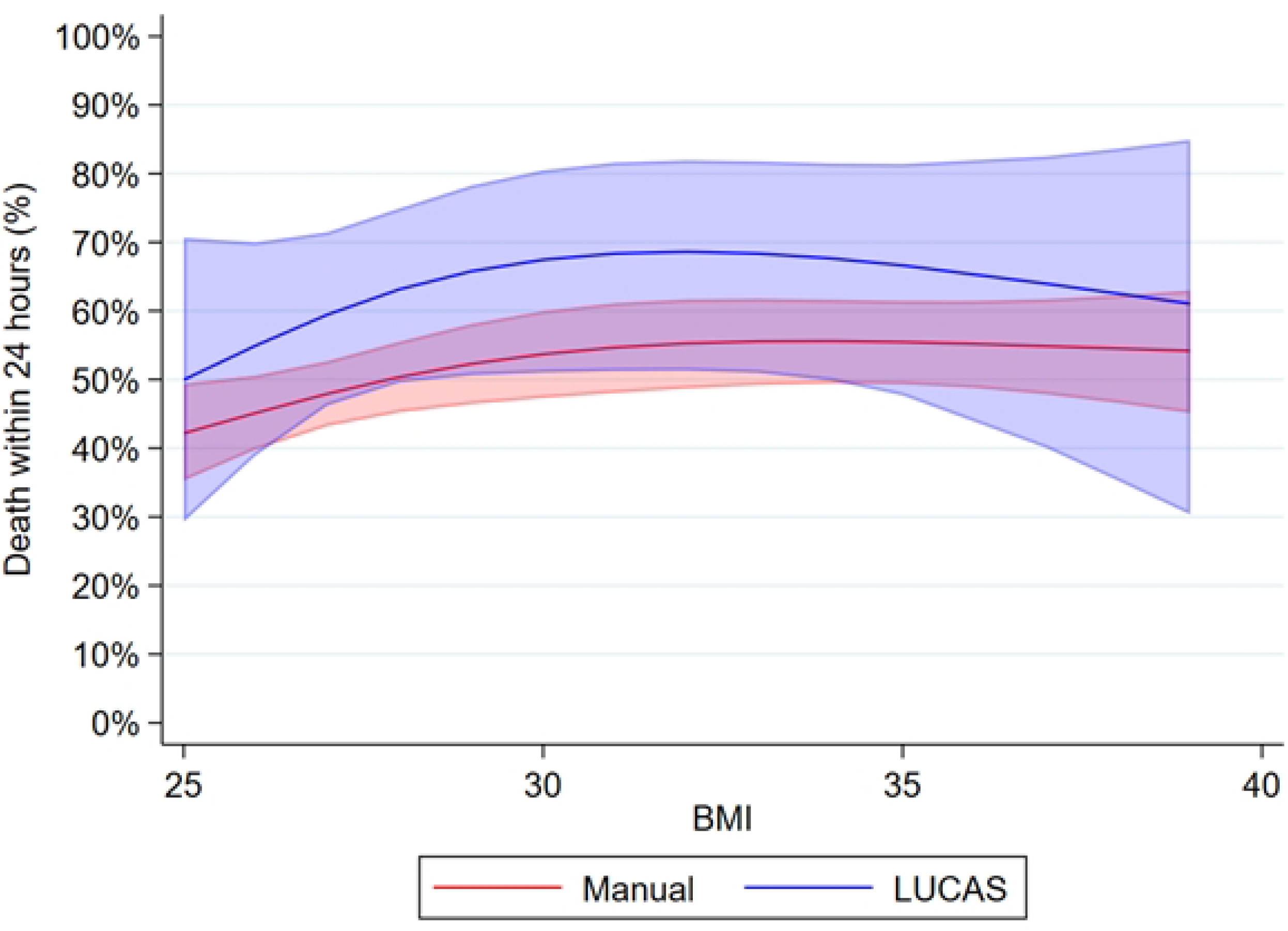
Predicted probability of death within 60 days after in-hospital cardiac arrest across the BMI range ≥ 25 kg m^-2^. Estimates are derived from the IPTW-weighted cubic-spline logistic model. The red curve represents manual CPR and the blue curve represents LUCAS® CPR; shaded areas denote 95% confidence intervals.

## Discussion

In this multicenter cohort of in-hospital cardiac arrests among overweight and obese patients, we found no significant difference in short-term or longer-term survival between LUCAS® mechanical CPR and high-quality manual chest compressions. Specifically, 24-hour mortality (our primary outcome) was similar with mechanical versus manual compressions, even after IPTW adjustment. Sixty-day in-hospital mortality was likewise not significantly different between groups. Importantly, no significant interaction was found between BMI and CPR methods, indicating that the effect of LUCAS was consistent across the overweight and obese spectrum.

Our finding of no difference in early survival aligns with several previous studies and meta-analyses evaluating mechanical versus manual CPR. A recent comprehensive meta-analysis of 24 studies (largely in out-of-hospital cardiac arrest) found no significant difference in short-term survival (defined as 24-hour survival) between mechanical and manual compressions [8]. The pooled odds ratio for 24-hour survival in that analysis slightly favored manual CPR (OR =0.77) but was not statistically significant, consistent with our point estimate suggesting no clear advantage to mechanical support. Large randomized trials in out-of-hospital settings have likewise reported equivalent early survival rates. For example, the LINC trial (using the LUCAS® device) showed nearly identical 4-hour survival between mechanical and manual CPR (=23.6% vs 23.7%) [2]. Similarly, in a scoping review of CPR in obese patients the variability in reported outcomes among obese adults indicates that obesity alone is not a factor to predict poorer prognosis. Therefore, the focus should be on delivering high-quality CPR and timely defibrillation, while recognizing the potential manual and mechanical challenges associated with resuscitation in obese patients [9]. Taken together, these external data reinforce our result that mechanical CPR does not substantially increase immediate survival after cardiac arrest. It is important to highlight that our study focused on an obese cohort (mean BMI ∼30 kg/m^2^), whereas most prior trials involved general populations with presumably lower average BMI. One might expect obesity to impair the quality of manual compressions due to reduced chest-wall compliance and rescuer fatigue, potentially giving mechanical devices an advantage. However, our results did not show any early survival benefit of LUCAS® even in this high-BMI group. A plausible explanation is that with well-trained responders (as in our hospitals), high-quality manual CPR can be maintained despite patient obesity, thereby mitigating any theoretical benefit of mechanical assistance. In other words, the challenge of achieving adequate depth in obese patients can often be overcome by skilled providers, so early outcomes remained comparable between manual and mechanical CPR across the BMI spectrum.

We also found no significant difference in longer-term outcomes (60-day in-hospital mortality) between mechanical and manual CPR. This finding mirrors the consistently null results for survival-to-discharge or 30-day survival reported in the systematic review [8], which reported a combined OR of 1.05 (95% CI: 0.84, 1.32) from three clinical trials [10–12]. Our data extend these observations to an in-hospital overweight and obese population. The lack of divergence in 60-day mortality curves between the LUCAS® and manual groups suggests that any advantages of mechanical CPR in generating perfusion did not translate into improved longer-term outcomes. It is important that increasing BMI in our cohort was associated with higher mortality risk up to a point (BMI ∼28 kg/m^2^) with a plateau thereafter, but this trend was essentially the same regardless of compression method. This finding is clinically relevant because although obesity can make manual CPR more challenging, current mechanical devices may have their own limitations in obese patients. For example, difficulties in application on a large torso or fixed compression depth may be insufficient for very thick chest walls. Furthermore, any potential hemodynamic benefit of uninterrupted, fatigue-free compressions by LUCAS® might be counterbalanced by issues such as deployment delays or suboptimal pad positioning in obese individuals.

This study has several strengths. It is among the first to evaluate mechanical CPR specifically in overweight and obese inhospital arrest patients, filling an important evidence gap as obesity rates rise. Our multicenter design across seven hospitals and a sizable cohort improves generalizability, and rigorous IPTW adjustment achieved excellent balance (all post-weighting SMDs < 0.1). Modeling BMI with splines allowed us to test whether LUCAS® effectiveness varied across the excessweight spectrum. Nonetheless, important limitations remain. As a retrospective, observational analysis, unmeasured confounding is possible, for example, clinicians may have selected LUCAS® for arrests with a poorer prognosis. Only 12% of patients received LUCAS® (n = 84), limiting power, and we lacked detailed CPR quality metrics (depth, fraction, pauses) that are critical mediators of outcome. We were not able to use waist circumference, a more reliable metric of obesity than BMI as it is not yet used as a vital sign in clinical practice [13]. We did not have waist circumference documented for all patients included in the study. We also did not assess neurological function or other morbidity, and our findings from wellresourced Mayo Clinic hospitals may not extend to settings with less CPR training or different workflows.

In this study we observed that implementing LUCAS® mechanical compressions for IHCA does not improve 24-hour or 60-day survival compared to manual CPR in patients with a BMI ≥ 25 kg/m^2^. Mortality rates in the critical early period and at hospital discharge were statistically equivalent between mechanical and manual compression groups across the overweight and obese spectrum. These findings suggest that in well-resourced settings with trained personnel, high-quality manual CPR may be sufficient even in obese patients, potentially reserving mechanical devices for specific logistical or staffing scenarios

## Conclusion

Our study underscores that among overweight and obese patients with in-hospital cardiac arrest, the LUCAS® mechanical CPR device did not improve survival at 24-hour or 60-day endpoints. Mechanical CPR also offered no incremental benefit across BMI categories, challenging assumptions of device superiority and highlighting the importance of high-quality manual CPR. These findings suggest outcomes are influenced by complex factors beyond the compression method, emphasizing the need for tailored resuscitation training and protocols for obese patients.

Future research should specifically address factors such as longer-term survival outcomes, real-time quality metrics of chest compressions, explore device modifications tailored to obese body habitus, and a prospective randomized approach to further clarify these interactions and enhance resuscitation strategies for obese populations.

## Data Availability

Data cannot be shared publicly because it contains Patient Health Information. Data are available from the Mayo Clinic Institutional Data Access / Ethics Committee for researchers who meet the criteria for access to confidential data.

## Acknowledgements

None

## Supporting Information

**S1 Fig**. Covariate balance before and after IPTW adjustment.

